# TEleRehabilitation Nepal (TERN) to improve quality of life of people with spinal cord injury and acquired brain injury. A proof-of-concept study

**DOI:** 10.1101/2021.06.21.21257001

**Authors:** Raju Dhakal, Mandira Baniya, Rosie M Solomon, Chanda Rana, Prajwal Ghimire, Ram Hariharan, Sophie G Makower, Wei Meng, Stephen Halpin, Shane Xie, Rory J O’Connor, Matthew J Allsop, Manoj Sivan

## Abstract

**Background:** Spinal Cord Injury (SCI) and Acquired Brain Injury (ABI) lead to unemployment, poverty, increased mortality, and decreased quality of life in low-and middle-income countries (LMICs). Telemedicine is increasingly facilitating access to healthcare, in LMICs. This prospective cohort intervention study aims to investigate feasibility and acceptability of telerehabilitation to provide long-term follow-up of individuals with SCI and ABI, in Nepal, post-discharge from hospital.

**Methods:** TERN was delivered by Spinal Injury Rehabilitation Centre, Nepal, in collaboration with University of Leeds, UK. A home visit connected participants to the Multidisciplinary Team (MDT), through a specialised video conference system. The MDT performed a comprehensive assessment before recommending personalised interventions. Outcome measures assessed functional independence in performing activities of daily living, health-related quality of life and emotional disturbances using Modified Barthel Index, EuroQoL-5D, and Depression, Anxiety, Stress Scale, respectively. A subset of participants was interviewed, exploring acceptability of telerehabilitation.

**Findings:** Between January and December 2020, 97 participants with SCI (*n* = 82) and ABI (*n* = 15) were enrolled. After receiving telerehabilitation, significant improvement to self-reported functional independence (*p*<.001) and quality of life were found, (*p*<.001). Self-reported severity of depression, anxiety and stress significantly decreased (*p*<.001). Qualitative interviews (*n* = 18) revealed participants accepted telerehabilitation, they valued regular contact with professionals without costly, difficult travel.

**Interpretation:** This is the first study to test telerehabilitation in Nepal. This approach can be safely delivered for long-term follow-up for individuals with SCI or ABI, overcoming geographical barriers to healthcare access. A larger-scale controlled study is required to further explore clinical and cost-effectiveness.

## INTRODUCTION

The disabilities an individual experiences after SCI and ABI (Acquired Brain Injury) often results in entering a spiral of unemployment, poverty, and ill-health.(1–3) Outcomes are particularly poor in low- and middle-income countries (LMIC) due to paucity of specialist rehabilitative services.(2) Despite rehabilitation having been included in the national health policy and planning in Nepal, no government hospitals are providing specialist rehabilitation services for the complex needs in this country with over 26 million people, out of which nearly 2% (513,132) are living with disabilities.(4) There are currently no established pathways for community-oriented rehabilitation for individuals with disability after discharge from hospital.(5) Travelling to hospital services are extremely challenging for both rural and urban dwellers with disabilities due to the mountainous terrains of the region and limited transport facilities. This group is known to experience multiple secondary complications, high mortality, and poor community reintegration.(6) Specifically, 1-2 years post-discharge from lengthy inpatient stay in hospitals, one quarter of people with SCI in Nepal die, rising to one third of those who used wheelchairs.(7) The vast majority have ‘severe’ or ‘extreme’ restrictions to community participation and one third of survivors are readmitted due to medical complications.(7) This contrasts the situation in several developed countries which have better survival and lesser complication rates due to good community rehabilitation services.

Mobile telecommunication connectivity has grown exponentially in Nepal. In 2020 the proportion of the population accessing broadband reached 80%, largely 3G and 4G mobile data services, whilst standard mobile telephone access has reached virtually the entire population.(8) Telemedicine is typically defined as the provision of health care services at a distance, which can include the use of information communication technology (ICT) for medical diagnostics, monitoring, and therapeutic purposes.(9) Broadly, there is emerging evidence for the effectiveness of telemedicine for improving outcomes for patients and its acceptability to both patients and health professionals. In Nepal, telemedicine (for diabetes management) and telephysiotherapy (for musculoskeletal problems) has been shown to be effective and cost saving in rural Nepal.(10,11) However, its application in supporting other disease groups, especially people living with long term physical and cognitive disabilities and remote delivery of rehabilitative care (also referred to as telerehabilitation) is limited.(12,13) There is a need to explore its feasibility, efficacy and cost-effectiveness in Nepal, particular given the low rehabilitation resources in the community and mountainous terrains of the country with limited transportation system. Whilst approaches to care delivery that utilise digital technology show promise in LMICs, an understanding of how these translate to address health system challenges and long-term rehabilitative care is yet to be explored well.

This study aimed to determine the feasibility and acceptability of a telerehabilitation approach for a cohort of individuals with SCI and ABI discharged from SIRC and estimate impact utilising outcome measures of function, disability and health.

## METHODS

### Setting and participants

SIRC, located near the capital city of Kathmandu, is a non-governmental organization (NGO) specialist multidisciplinary rehabilitation centre for people with SCI and ABI. The services offered at SIRC includes medical care, nursing, physiotherapy, occupational therapy, psychology, peer counselling, vocational training, social service, community-based rehabilitation, prosthesis, orthosis and assistive devices, and other extended services (such as speech therapy and recreational therapies). The TERN project was developed by collaboration between SIRC and the Academic Department of Rehabilitation Medicine, University of Leeds, UK.

A prospective, within subject, uncontrolled study was conducted to measure the impact of telerehabilitation on the outcomes of people with SCI and ABI who were unable to access the centre for follow-up care after discharge.

Inclusion criteria

- Age 18 years or above
- Diagnosis of SCI or ABI and received inpatient care in SIRC
- Discharged from SIRC between February 2018 and August 2019.

Exclusion criteria: Individuals who do not report any ongoing rehabilitation needs

Demographic characteristics captured were age, sex, distance from SIRC, terrain, nature of disability, duration since injury, history of consultation at other hospitals since discharge, employment, marital status before and after disability, and ability to leave house.

### Outcome measures

#### Modified Barthel Index (MBI)

MBI measures performance of an individual items of activities of daily living (ADL) i.e., feeding, grooming, bathing, dressing, continence of bowel and bladder, transfer to and from wheelchair, transferring to and from toilet, use of wheelchair, use of stairs, and walking. The items are scored based on the amount of physical assistance required to perform the task. Each item has 5 categories, the first category indicates dependence, and the fifth category indicates independence. The total score ranges from 0 to 100 where a higher score indicates increased independence in performing ADLs.(14) The tool was found to be valid and reliable for SCI population. Internal consistency of MBI was reported to be 0.88 in a previous study.(15)

#### Depression Anxiety Stress Scale (DASS)

DASS-21 is a self-report measure derived from 42 item DASS scale. DASS-21 consists of three 7-item subscales: depression, anxiety, and stress. The items refer to the feelings that occurred past week. Each item is scored on a 4-point scale (0 = “did not apply to me at all”, to 3 = “applied to me very much or most of the time”). The score for each subscale ranges from 0 to 21, where higher score indicates greater severity. DASS-21 is a valid, reliable, and easy to use tool; the total scores range between 0 to 63.(16)

#### EuroQoL 5 (EQ-5D-5L)

EQ-5D-5L was used as a measure of assessing health related quality of life (HRQoL) of the participants. It is a 5-level version of EuroQoL. It comprises of 5 dimensions of health: mobility, self-care, usual activities, pain/discomfort, and anxiety/depression; each dimension is scored on a 5-point scale (1 = “no problem” to 5= “unable to /extreme problem”). In addition, the visual analogue score (EQ VAS) was used to measure the direct valuation of the current state of health of participants on a 0-100 scale, where ‘0’ refers to “the worst health you can imagine” and 100 refers to “the best health you can imagine”. This tool exhibited excellent psychometric properties amongst the SCI and ABI population.(17) EQ-5D index value was derived from the five dimensions of EQ-5D-5L and was calculated using the EQ-5D-5L Crosswalk Index Value Calculator.(18)The EQ-5D index value ranges from 0 to 1, where 0 indicates severely ill, and 1 indicates a perfect health.

A 5-point Likert scale was used to assess the perception participants about benefit of telerehabilitation which ranged from ‘1’ completely disagree to ‘5’ completely agree that telerehabilitation was beneficial.

This study was not powered to detect clinically significant changes in function, disability and health as this was primarily a feasibility study. Multiple validated outcome measures were used for baseline and post-intervention assessment of the participants to estimate an effect size that could be used in a larger efficacy evaluation trial in future. The outcome measures were available in both English and Nepali languages and participants could choose the language they were comfortable with. For those with cognitive difficulties, primary caregivers or family members were involved in the data collection.

#### Sampling and data collection procedure

A list of 129 consecutively discharged participants between February 2018 and August 2019 were contacted by telephone to discuss the study and obtain verbal consent if they were suitable and willing to participate. Study protocol and consent form were sent to participants by post. After gaining verbal consent, a home visit by a member (social worker) of the SIRC team was planned for each participant. On the home visit, the social worker explained the nature of the study and obtained written consent. Following a baseline assessment, the social worker connected the participant via a mobile phone device to SIRC via the telerehabilitation system at SIRC. The telerehabilitation team at SIRC comprised of an MDT of six members including a rehabilitation physician, rehabilitation nurse, physiotherapist, occupational therapist, and social worker. As part of the project, specialized audio-visual device (smart led television, People link UVC, People link Quordo, People i Com WHD camera) and internet system was installed in the MDT room to facilitate the remote consultation. The telerehabilitation consultation was provided via InstaVC, a video conferencing platform. In cases where InstaVC could not be used, social media platforms or audio conferencing, via telephones, were used. The social media platforms used included Facebook messenger, WhatsApp and Viber™. All consultations took place between January and December 2020.

#### Telerehabilitation interventions

The social worker assisted the participant to video conference the MDT for the first telerehabilitation consultation. The participant was remotely examined. New and existing health problems, in relation to the participant’s SCI, were assessed. The team then provided management and treatment, where necessary.

During an initial telerehabilitation session, the MDT discussed the ongoing physical, cognitive, psychological and vocational problems the individual was experiencing. The problems which could be intervened at home using local resources were immediately addressed during the consultation by the team. This included use of medications, advice on skin care, catheter care, exercises, use of assistive and mobility aids, dietary plans, and counselling on general coping skills (Table 5). Equipment including walking aids, catheters, mattresses or hospital bed were arranged to be delivered in selective cases. There was the potential to refer participants to the nearest hospital for specific diagnostics and treatment or to be admitted for inpatient rehabilitation at SIRC. Some participants needed more than one follow-up assessment. During these remote appointments, goals were reviewed, and outcome measures completed. The second follow-up telerehabilitation was delivered between 1-7 weeks later, depending on the treatment plan derived in the first consultation. After completing both telerehabilitation consultations, the post-intervention assessment was performed.

#### Qualitative interviews

A subset of participants (*n*= 18) was purposively selected across variables including participants’ sex, rural or urban residential location, and type of disability. In-depth qualitative interviews were conducted to explore participant experiences and acceptability of the telerehabilitation intervention.

### Data analysis

Descriptive statistics was used to describe demographical and injury related characteristics of the participants. Binary and categorical data are presented in frequencies and percentages.

Continuous variables are presented as means with standard deviation (SD). Normality of the dependent variables was determined assessing skewness, kurtosis, and histogram. Paired sample t-test or Wilcoxon signed rank test were applied to compare the pre and post-test data.

Significance level was considered at p value < 0.05. Cohen’s d effect sizes were used. Data was analyzed using SPSS (SPSS: Version 20.0. Chigago, IL, USA). Thematic analysis was applied to inductively identify emergent themes from the qualitative data. All transcripts were independently coded and organised into potential themes. NVivo software was used to arrange the qualitative data.

### Ethical consideration

Ethical approval for the study were obtained from the Nepal Health Research Council (reference: 1727) and the University of Leeds School of Medicine Research Ethics Committee (reference;MREC 19-031). This study is registered under clinicaltrials.gov (ClinicalTrials.gov Identifier: NCT04914650).

## RESULTS

Among 129 participants approached, 97 were eventually included in the study. Figure 1 displays the flow of participants.

**Figure 1.**
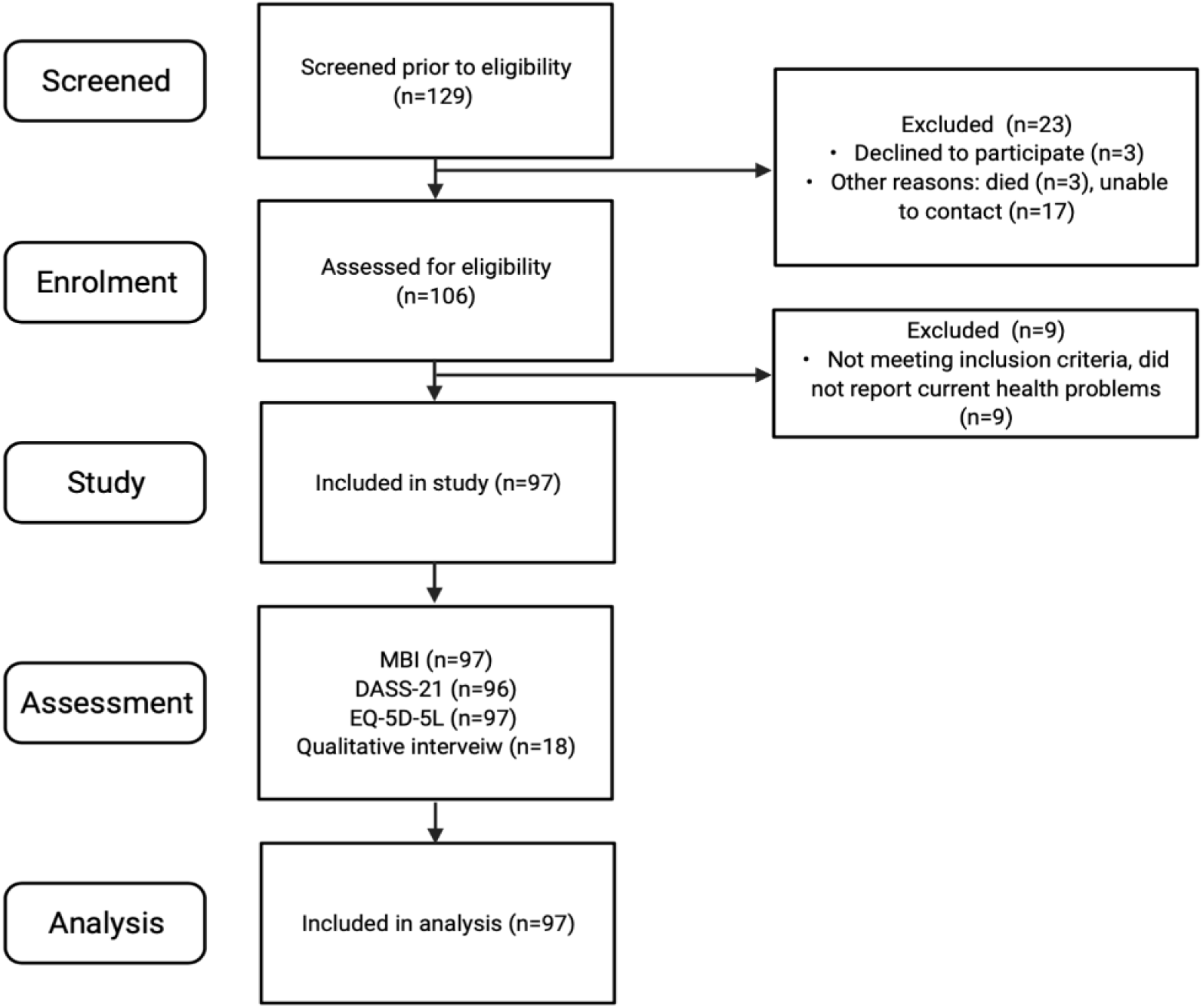
Flowchart of number of participants approached and included

**Figure 2.**
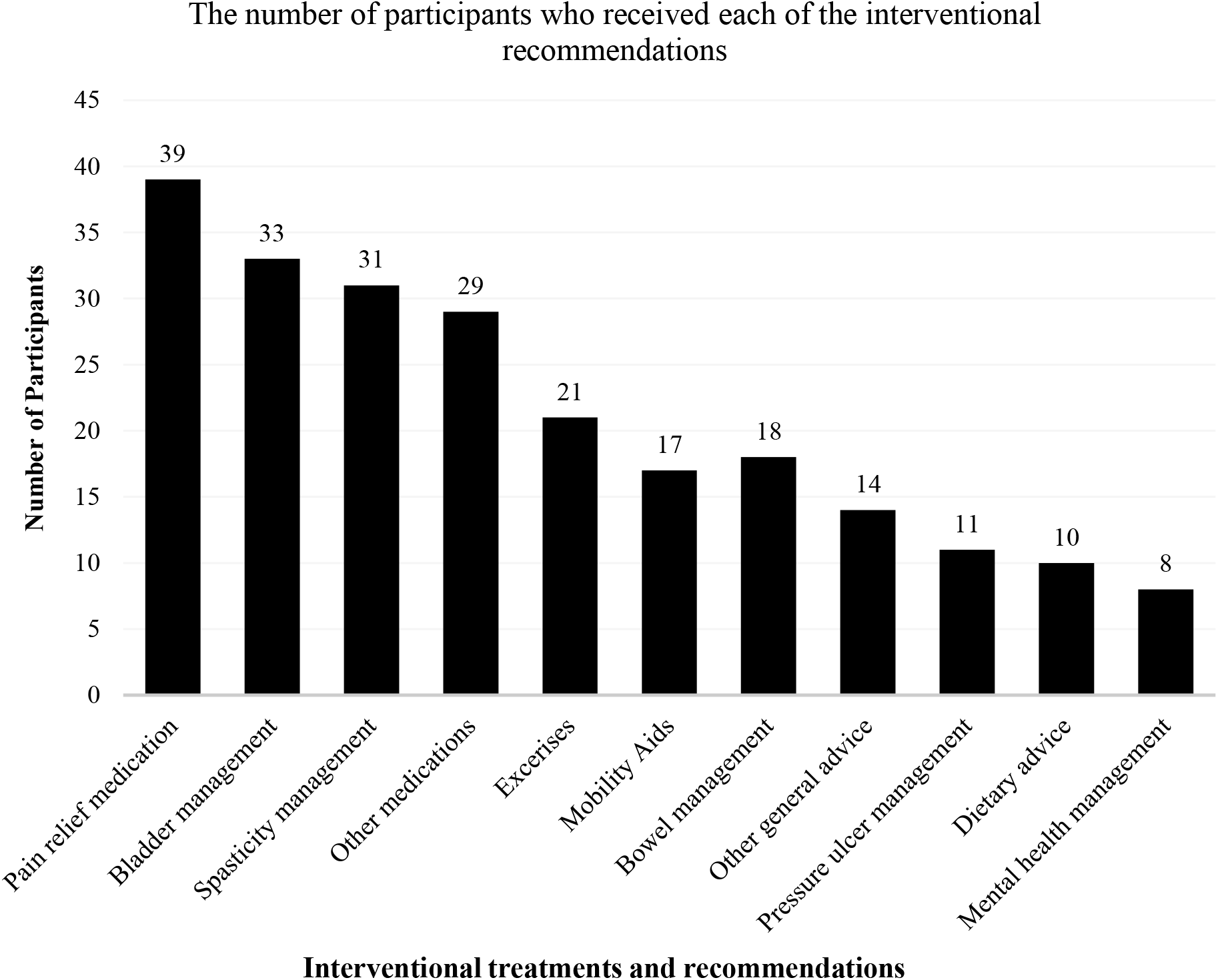
Interventional treatments and recommendations

The mean (SD) age of participants was 38.4 (13.2) years (range 18 to 73 years). More than one-third was male (79.5%) and had SCI (84.5%). Nearly 60% participants were from hilly regions. Eighty-six percent of participants were employed before injury whereas after injury only 13% managed to retain their employment. Less than half (43.3%) participants were able to leave their house without assistance at first assessment. (Table 1)

**Table 1.**
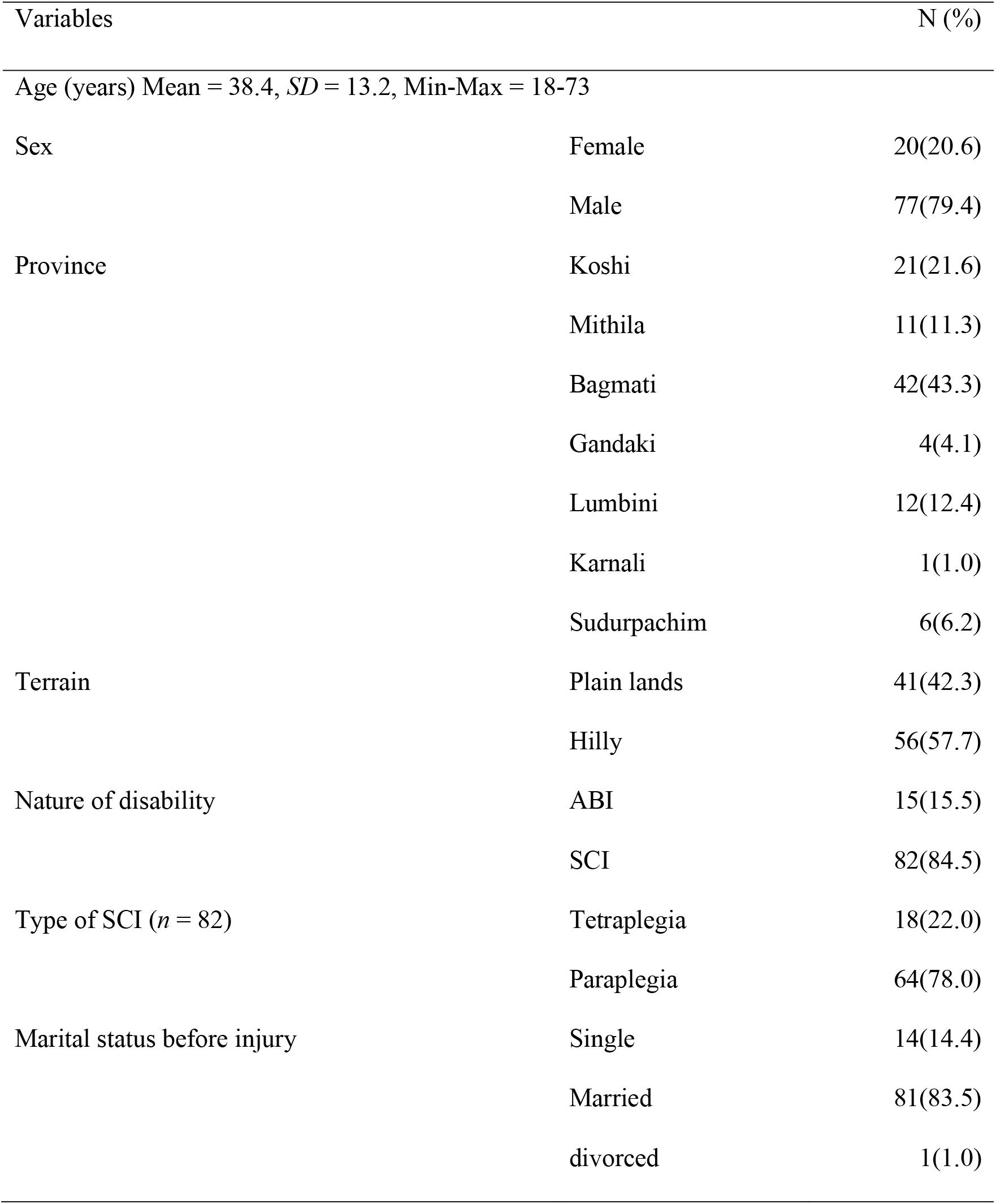
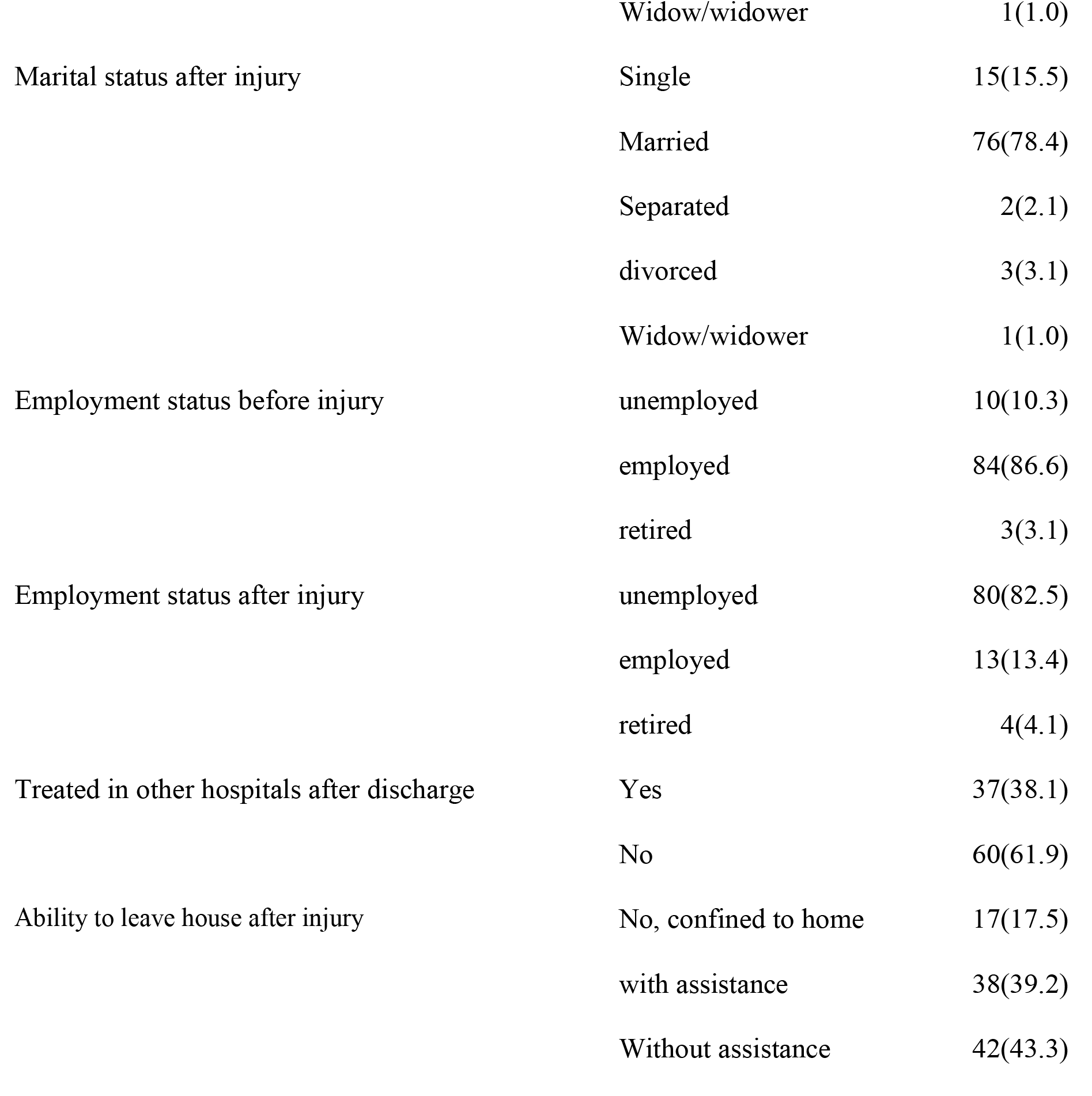
Demographic and disability related characteristics of the participants (*n*=97)

The severity of depression, anxiety and stress significantly decreased after intervention (*p*<0.01). The EQ-5D index score significantly increased in post intervention with *p* < .001 (Wilcoxon signed rank test) (Table 2). Before receiving telerehabilitation, 10(10.3%) participants completely agreed (Likert scale) that telerehabilitation intervention will be beneficial for them which increased to 17(17.5%) after the intervention.

**Table 2.** Pre and post-intervention DASS and EQ-5D-5L domain scores

| Variables | Pre-intervention n(%) | Post-intervention n(%) | *p* -value† |
| --- | --- | --- | --- |
| Depression (*n*=96) |  |  | <.001 |
| Normal | 66(68.8) | 68(70.1) |  |
| Mild | 6(6.3) | 10(10.3) |  |
| Moderate | 16(16.7) | 13(13.4) |  |
| Severe | 3(3.1) | - |  |
| Extremely severe | 5(5.2) | 5(5.2) |  |
| Anxiety (*n*=96) |  |  | .001 |
| Normal | 87(90.6) | 91(94.8) |  |
| Mild | 5(5.2) | 2(2.1) |  |
| Moderate | 2(2.1) | 1(1.0) |  |
| Severe | - | - |  |
| Extremely severe | 2(2.1) | 2(2.1) |  |
| Stress (*n*=96) |  |  | <.001 |
| Normal | 77(80.2) | 85(88.5) |  |
| Mild | 10(10.4) | 4(4.2) |  |
| Moderate | 5(5.2) | 4(4.2) |  |
| Severe | 2(2.1) | 1(1.0) |  |
| Extremely severe | 2(2.1) | 2(2.1) |  |
| EQ-5D-5L domains |  |  |  |
| Mobility |  |  | .006 |
| No problem | 32(33.0) | 36(37.1) |  |
| Slight problem | 34(35.1) | 35(36.1) |  |
| Moderate problem | 15(15.5) | 13(13.4) |  |
| Severe problem | 5(5.2) | 5(5.2) |  |
| Unable to walk | 11(11.3) | 8(8.2) |  |
| Self-care |  |  | <.001 |
| No problem | 32(33.0) | 36(37.1) |  |
| Slight problem | 12(12.4) | 12(12.4) |  |
| Moderate problem | 23(23.7) | 25(25.8) |  |
| Severe problem | 19(19.6) | 14(14.4) |  |
| Unable | 11(11.3) | 10(10.3) |  |
| Usual activities |  |  | .004 |
| No problem | 44(45.4) | 49(50.5) |  |
| Slight problem | 29(29.9) | 26(26.8) |  |
| Moderate problem | 9(9.3) | 9(9.3) |  |
| Severe problem | 8(8.2) | 7(7.2) |  |
| Unable | 7(7.2) | 6(6.2) |  |
| Pain/discomfort |  |  | <.001 |
| No pain | 14(14.4) | 15(15.5) |  |
| Slight pain | 28(28.9) | 47(48.5) |  |
| Moderate pain | 24(24.7) | 31(32.0) |  |
| Severe pain | 24(24.7) | 4(4.1) |  |
| Extreme pain | 7(7.2) | - |  |
| Anxiety/depression |  |  | <.001 |
| Not anxious | 27(27.8) | 33(34.0) |  |
| Slightly anxious | 35(36.1) | 39(40.2) |  |
| Moderately anxious | 17(17.5) | 17(17.5) |  |
| Severely anxious | 16(16.5) | 6(6.2) |  |
| Extremely anxious | 2(2.1) | 2(2.1) |  |

†Wilcoxon signed rank test; MBI = Modified Barthel Index (SHAH version); DASS-21 =
Depression, Anxiety, Stress Scale 21; EQ-5D-5L = EuroQol 5 Domains

There was a significant mean difference (*p*<.001) between the pre- and post-intervention MBI and EQ VAS score with effect size -0.4 and -0.7 respectively (Table 3).

**Table 3.** Mean difference between pre and post-test MBI scores EQ VAS score

| Variables |  | Mean | SD | Mean | *p*- | Cohen's d | 95% CI of B |  |
| --- | --- | --- | --- | --- | --- | --- | --- | --- |
|  |  |  |  | difference | value* | Effect size | Lower | Lower |
| MBI Total | Pre-intervention | 60.9 | 27.3 | -1.5 | <.001 | -0.4 | -.60 | -.19 |
|  | Post-intervention | 62.4 | 27.3 |  |  |  |  |  |
| EQ VAS | Pre-intervention | 60.0 | 25.7 | -5.7 | <.001 | -0.7 | -.92 | -.47 |
|  | Post-intervention | 65.6 | 22.3 |  |  |  |  |  |

\* Paired sample *t*-test

**Table 4.** Findings from qualitative interview

| Topic | Summary | Supporting quotes |
| --- | --- | --- |
| Acceptability | Participants were receptive to the concept of telerehabilitation. Most participants lived a long distance from the Centre. Participants reported that travelling by typically inaccessible public transport is burdensome and creates hassle which can be avoided with telerehabilitation.<br>Even in urban areas, participants reported that it is difficult to access health services for people with SCI and ABI. They considered telerehabilitation as a useful means of accessing health services during emergencies from home. | *“This is extremely important step taken because I feel being a wheelchair user, with poor accessibility we cannot even access the basic health needs. In this condition to be able to connect back to SIRC team will ease so many hassles of transportation and to be able to access tele consultation from home will ease our life to great extent.”* Male, SCI, urban community<br>*“It is a very good idea. I am very positive about it. Although we live in city area, we do not know many things about the management of various health problems (for her father). It must be more difficult in rural areas. For emergency help it is very helpful. You have started a very good program. I appreciate this* |
|  |  | *initiative.” Caregiver (of participant with stroke), female, urban community* |
| Recommendation | Participants from urban areas requested planned online meetings to accommodate family members who had jobs which could be done by selecting a fixed topic for discussion for a planned day where participants were keen on the ability to be able to present their problems and those of their caregivers in real-time to the rehabilitation team. | *“I think it would be better to have a specific day, time, and specific topic for discussion as part of telerehabilitation program that you are going to start. If there is a team that is available may be on Saturday or Sunday afternoon for an hour with a pre informed topic for discussion, we can arrange our time to attend the tele meeting and discuss our problems.” Caregiver (of participant with stroke), female, urban community* |

**Table 5.** Interventional recommendations provided during telerehabilitation

| Type of intervention | Number of participants | Details |
| --- | --- | --- |
| Pain relief | 41 | Medication: Anticonvulsants- Pregabalin ($n=37$), Gabapentin ($n=1$); NSAIDs- Aceclofenac ($n=12$), Etoricoxib ($n=2$), Tricyclic Amitriptyline ($n=11$)<br>Physical modalities (Heat pack) ($n=1$) |
| Exercises | 21 | Progressive resistance n=and passive range of motion exercises for upper and lower limbs ($n=26$), back ($n=3$), pelvic floor ($n=4$)<br>Passive range of motion ($n=5$), Tenodesis grip ($n=1$) |
| Spasticity management | 31 | Baclofen ($n=31$), Tizanidine ($n=13$) |
| Other medications | 30 | Proton pump inhibitors- Omeprazole ($n=28$), H2 blocker-Ranitidine ($n=1$)<br>Antibiotics- Amoxyclav ($n=1$), Cefixime ($n=1$),<br>Calcium, Vitamin D3 and Vitamin B12 ($n=21$)<br>Zolpidem ($n=1$), Rotacap Salmeterol and Fluticasone inhaler ($n=1$)<br>Anti-emetic-Domepridone ($n=1$) |
| Assistive products | | Commode chair ($n=9$), modified spoon($n=1$) |
| ADL aids |  |  |
| Mobility aids | 17 | Ankle Foot Orthosis (*n*=3), Wheelchair (*n*=2), Knee brace orthosis (*n*=1), Walker (*n*=1) |
| Bladder management | 33 | Medication: Anticholinergics-Tolterodine (*n*=18), Oxybutynin (*n*=1)<br>Antibiotics (*n*=1), Tricyclic antidepressant (*n*=2), Urine alkalyzer (*n*=2)<br>Investigations (urine routine and microscopy *n*=2, ultrasound kidney, ureter, bladder *n*=2)<br>Bladder catheterization- Clean intermittent self-catherization *n*=10, indwelling *n*=1; Bladder diary (*n*=5) |
| Bowel management | 19 | Medication- Bisacodyl (*n*=10), lactulose (*n*=6), Cremaffin syrup (*n*=5); Herbal ointment (*n*=5) for haemorrhoids<br>Bowel training advice (*n*=5), bed pan (*n*=1) |
| Pressure ulcer | 11 | Pressure relief positioning (*n*=9), wound dressing (*n*=11) |
| Dietary advice | 11 | High protein diet (*n*=6), high fibre diet (*n*=3), low calorie diet (*n*=2), general dietary advice (*n*=2) |
| Mental health management | 8 | Medication- Tricyclic anti-depressant-Amitriptyline (*n*=4), Beta-blocker-Propanolol (*n*=1)<br>Psychological counselling (*n*=1) |
| Sexual and reproductive advice |  | counselling (*n*=2), medication- Sildenafil (*n*=1) |
| Other general advice | 14 | ADLs and self-care advice: transfer (*n*=5), dressing (*n*=2), bathing (*n*=1)<br>Gait evaluation and advice (*n*=3)<br>Referral- Ophthalmology (*n*=1), X-ray (*n*=2) |

Note: \*One participant was prescribed one or more than one intervention

### Adverse effects

Eleven participants had pressure ulcers at the first telerehabilitation consultation, all were provided with advice including lying prone and redressing the ulcer daily with Betadine. Two participants had to stop taking Pregabalin, which was prescribed to manage their neuropathic pain, because it made them too drowsy. One participant experienced increased pain lying in the prone position because of fractured ribs. One participant sat in an unrecommended position in his wheelchair. When the team made recommendations for his sitting position, he found it more uncomfortable but relieved the pain by returning to his original sitting position.

## DISCUSSION

This study assessed the feasibility of a telerehabilitation approach for following up and providing interventions to those with physical, cognitive, and psychological problems from SCI or ABI in Nepal. This study finding suggests that telerehabilitation intervention may increase independence in carrying out ADLs and improve psychological health and quality of life among people with ABI and SCI. In addition, most participants in qualitative interview reported telerehabilitation was beneficial to address their health needs and welcomed the new approach. Our findings support the use of telerehabilitation to improve the access to rehabilitation services. The main benefit of telerehabilitation is being able to reach out and providing support to those living in remote area isolated from access to basic health services. However, literature shows inconsistent findings.

A recent systematic review of randomized control trials (RCTs) that included forty-eight trials found that high and moderate quality evidence demonstrated telerehabilitation was not different to other interventions for physical problems, functioning and quality of life. (19) However, some low-quality evidence and a small number of trials found significant differences in telerehabilitation group compared to control group. In some RCTs, telerehabilitation showed significant improvement in balance and mobility (*p* =.001), (20) MBI scores, balance (*p*<.001)(21) among stroke survivors. Likewise, in another RCT, participants with SCI received 1 hour counseling per week for 8 weeks for leisure time physical activity (LTPA) motivation. The intervention group reported greater autonomous motivation (Hedge’s *g* = 0.91) and LTPA (Hedge’s *g* = 0.85) post-intervention. (22) In a Nepalese feasibility study of telephysiotherapy, delivered for four weeks, there was a significant reduction in various musculoskeletal pain (used NPRS) (at rest, during ADL, during occupation).(11) In another study, trained peer trainer provided telephone based intervention for six months that resulted in improvement in self-management for prevention of secondary complications, increased service use (estimate, 1.54; *p*=.007) increased life satisfaction (estimate, 1.00; *p*=.052),among adults with SCI. (23)

Various studies demonstrated telerehabilitation was effective for psychological health. A systematic review concluded that telephonic counselling was more effective compared to face-to-face counselling to reduce depressive symptoms.(24) In a RCT, 12 weeks physical activity counseling training was delivered by telephone which significantly increased physical activity and decreased severity of depression among people with multiple sclerosis.(25) Similarly, other RCTs found that cognitive behavior therapy delivered through telephone for 8 to 12 weeks significantly decreased patient reported symptoms (treatment effect = 0.36, 95% CI: 0.01–0.70; *p* = 0.043)(26) and decreased depression scores significantly at 6 months follow up (*β* =-.41, *p*=.018).(27)

A meta-analysis showed that telemedicine was significantly effective to increase QoL among lung cancer patients [standard mean difference= 0.96, 95%, CI: 0.29–1.63].(28) Another study used a 12-Item Health Survey scale among veterans who were provided home based video physiotherapy that showed significant improvement in QoL after intervention (*p* = 0.02, *r* = 0.42). All participant veterans also reported satisfaction with their telerehabilitation experiences.(29)

In Nepal, the current post-discharge rehabilitation care for individuals with SCI and ABI is not meeting individuals’ needs. There are a lack of qualified rehabilitation service providers and the cost and burden of transportation for individuals with physical disabilities is high. Individuals have ongoing health problems, such as bladder/bowel issues, pain, spasticity, pressure injuries, and psychological problems. The reality is that majority of people with SCI and ABI, in Nepal, are discharged directly to their homes from after acute management with minimal advice about home exercises. In the context of Nepal, only a small number of ABI and SCI survivors are able to access rehabilitation services. Barriers for this include resource limitation, geographical barriers, time constraints, and lack of awareness about rehabilitation services. Perhaps telerehabilitation is the only way to provide follow-up, as is addresses at least some of the unmet rehabilitation needs. Telerehabilitation interventions are expected to decrease the symptoms of people with SCI and ABI and decrease some burden of care on caregivers.(30)

Various types and duration of intervention were delivered through telerehabilitation in the past studies. In our study, we delivered need-based interventions, and advises where there was no fixed duration of coaching for each intervention provided, however, the telerehabilitation mostly addressed most common issues related to bowel/bladder, pain, exercises, and nutrition. To improve effectiveness of a telehealth programs, interventions should focus on most common problems such as deconditioning, lack of nutrition, pressure point care, bowel/bladder care and pain management.(23) In LMIC settings, the telerehabilitation interventions should be simple, robust, and user-friendly for easy operability by less sophisticated technology and with the resources that are locally available. In addition, including a family member in a telerehabilitation could assist in increasing adherence of the intervention provided.(30) Moreover, there is still lack of uniformity in the intervention delivered virtually in terms of type, duration of intervention and follow up, and outcome measures used.(13)

The coronavirus disease 19 (COVID-19) pandemic caused a pause in our ongoing data collection. However, telerehabilitation proved useful as people could be seen and managed remotely. Telehealth is a beneficial alternative in the time of COVID-19 pandemic.(31) Our study has demonstrated that telerehabilitation has benefitted patients during the current Covid-19 pandemic. Although the internet and smart phone usage has rocketed in Nepal in the recent years, still only 34 % of this study participant owned a smart phone. A reliable internet connection was another challenge. Internet data had better connectivity; however it was very costly and most participants couldn’t afford it. In addition, some problems were encountered while using video demonstrations for intervention such as exercises and transfer techniques. It was realized that, despite the use of a specialized video conferencing system, it would be more convenient to use pre-filmed videos for common topics. This was easier for patients to watch and understand and saved time during consultations, so that other issues could be addressed. Adherence to the prescribed intervention was a problem for some participants. The reasons include (i) caregiver had difficulty in assisting to complete the intervention (the only family member was unwell and could not perform catheterization), (ii) they were unwilling to carry out requested interventions, (iii) prescribed medication was either not available in the local pharmacy or the participant did not want to take medicines because reportedly it was not effective to reduce pain and/or spasticity.

Ministry of Health and Population (MoHP), Government of Nepal developed a national e-health strategy 2017 with a vision to facilitate equitable and high-quality health care services to enable all Nepali citizens to enjoy productive and quality lives. Information and Communication Technology as a part of e-health can have a significant role in promotive, preventive, curative, rehabilitative and palliative health care services in the context of Nepal. This study used telerehabilitation to reach to the participants with physical disability (ABI and SCI) living at a range of 15 to 200 kilometres away from the specialist rehabilitation facility who would otherwise be abstained from rehabilitation services.

We could assume that initiation of this telerehabilitation program was helpful to facilitate relationships between clinicians and patients, and to motivate and activate patients to continue with the rehabilitation process at home. The literature demonstrates the possibility to collaborate among the providers so that in a situation where there is scarcity of rehabilitation professionals, there is a possibility of connecting to national and international professionals to provide telerehabilitation.(32) Furthermore, this study has flagged up the need for a specific criterion forming decision making for deciding interventions in future which could possibly improve adherence. A systematic and structured evidence-based guideline should be developed for telerehabilitation intervention. This study enhanced collaboration among health professionals and enabled them to actively participate and explore their skills and in diagnosis and decision-making. The findings of this study can be used to inform training programs for the rehabilitation professionals to develop skills and the provision of effective delivery of telerehabilitation intervention.

The study has several limitations. The sample size was small, observed changes to outcome may not be powered. There was no control arm to compare the treatment to. Participants were not followed-up long-term, a future study needs to identify such effects. Many outcomes were self-reported, which may have introduced response bias, especially since the participants were unblinded to the nature of the study. Despite such limitations, the study is sufficient to demonstrate feasibility of telerehabilitation and the positive attitudes towards its use.

Given the acceptability and feasibility of the telerehabilitation interventions seen in this study, there is need for further larger controlled studies to accurately capture the efficacy of this new approach of rehabilitation in Nepal and other similar low-resource countries. We also need to enrol more participants with cognitive impairments, to explore whether such impairments challenge remote care delivery. Cost-effectiveness evaluations of telerehabilitation approaches must be performed to as influence health policies in Nepal and other LMICs. Given the benefits seen in this study, there is also a need for reverse translation studies in high resource countries as these interventions using simple technology can lead to prompt and effective care being delivered and lead to additional savings in developed countries too.

## CONCLUSION

This is the first study in Nepal to show that telerehabilitation can be used to provide follow-up care to individuals with SCI or ABI. This study provides some preliminary evidence on efficacy of the benefits of telerehabilitation for these individuals. Telerehabilitation interventions were delivered effectively and they overcame geographical barriers to healthcare access. There were no observed side-effects or risks in this approach. A larger controlled trial is needed to demonstrate clinical and cost-effectiveness.

## Data Availability

All data referred to in this manuscript are available.

## ACKNOWLEDGEMENTS

We would like to thank all administrative staff at SIRC for their efficient organisation of the project with special thanks to Hari Adhikari, SIRC Director. We would also like to thank Esha Thapa and rest of the board members of the Spinal Injury Sangh Nepal (SISN) charity for their collaborative working with University of Leeds research team. This project would not have been possible without the cooperation and patience of all the participants in Nepal and their families and carers, and they remain our inspiration. Angela Greenbank at University of Leeds organized the travel of University of Leeds research team to Nepal with utmost care and attention to detail.

